# A Comparative Study for Blockchain Applications in Nursing Informatics

**DOI:** 10.1101/2024.02.24.24301619

**Authors:** Haeun Kim, Claire Lee, Diya Pendyala, Angela Ng, Tsung-Ting Kuo

## Abstract

We explored blockchain’s applications in nursing informatics, highlighting its potential to improve patient care and data management. We compared and analyzed eight studies focusing on blockchain in Electronic Health Records (EHR) management, nursing optimization, and research facilitation. Although most of these studies are in the proposal stage, blockchain’s technical features show promise in enhancing nursing practices and supporting nursing informatics researchers with the integration of technologies.

## Introduction

Nursing informatics systems are critical for the daily duties of nurses, who constitute the largest occupation in hospitals, with over 1.8 million jobs or 30 percent of total hospital employment^1^. For example, nurses use nursing informatics systems to collect and utilize patients’ data for critical tasks such as calculating important patient care metrics (length of stay, patient acuity/dependency), budget planning, and bed turnover management. In addition, nurse managers use nursing informatics databases to strategically adjust the planned nurse staffing levels based upon the trends of patient care. However, centralized data storage poses security risks and availability vulnerability, such as single-point-of-failure. Furthermore, the stored data are mutable, meaning they are susceptible to accidental changes of records. To mitigate these challenges, blockchain technology, with its decentralized, and immutable features, can be beneficial for Electronic Health Record (EHR) and medical informatics systems^2^. Nevertheless, there is yet to be a comparative analysis of nursing blockchain studies that provides insights into nursing informatics application areas, blockchain adoption status, and the integration of other technologies.

## Methods

This study aims to deepen the understanding of potential nursing blockchain use cases by comparing and analyzing recently published nursing blockchain literature. We adapted the Preferred Reporting Items for Systematic review and Meta-Analysis (PRISMA) approach^3^. Since PRISMA was originally designed to summarize clinical studies, we instead focused on extracting the following 13 data items: title, published year, journal name, source of publication, authors, countries of authors, nursing application, research focus, major technologies, blockchain adoption status, blockchain platform, and data/code availability. Our literature search encompassed two academic search engines, PubMed and Google Scholar, and identified studies published by November 2022. We used the phrase “*blockchain nursing”* as our initial search term and included all publications in the English language that discussed blockchain applications to the nursing field. Next, we removed all duplicated articles and manually filtered relevant ones. Titles, abstracts and full articles were subsequently screened by HK, with discussion to TTK, using eligibility criteria to exclude nursing education studies. Afterwards, all team members (HK, CL, DP, AN, and TTK) reviewed the selected papers, and the data items were extracted from the eligible studies.

## Results

Our study identified 8 publications (as shown in **Table 1**) and summarized our key findings in **Figure 1**. In general, blockchain applications in nursing include Electronic Health Records (EHR) management for nursing informatics, nursing management optimization, and facilitation of nursing research processes. Specifically for electronic nursing records, blockchain can enhance its control/usage, optimize nursing management, and ensure secure patient data ownership for nursing research. These publications span from 2018 to 2022, with research conducted in four countries. American (37.5%) and Chinese (37.5%) research institutes published the most studies. The distribution reveals that the United States and Australia had more publications between 2018 to 2020, while China and Hong Kong conducted more studies in 2021 and 2022. We categorized these eight papers into five blockchain adoption statuses: proposed, designed, developed, tested, and deployed. Four papers were considered as “proposed”, and provided introductions to blockchain technology, its characteristics, and its potential applications in nursing practice. Three “designed” papers offered more detailed insights into adopting for the specific units (e.g., obstetrics, nursing homes and nursing research). The only “tested” paper developed and tested a blockchain platform for nursing record management. We also found that several studies mentioned the integration of blockchain with other technologies to address various nursing informatics needs. For example, smart medicine^5, 8^ utilizes big data and blockchain for nursing information transfer to improve patient care and address data-related challenges. Another example is the nursing Internet of Things (IoT) system^6^, which can benefit from blockchain integration for efficient care service monitoring and healthcare task scheduling by using a genetic algorithm. Finally, nursing cloud storage^8^ combined with blockchain can offer enhanced data security and protection against insider threats.

**Table 1.** The list of 8 publications identified in our study.

| Authors | Year | Venue | Title |
| --- | --- | --- | --- |
| Carroll | 2020 | Nursing | How blockchain can improve nursing<sup>4</sup> |
| Sun et al. | 2021 | J. Health. Eng. | Obstetrics nursing and medical health system based on blockchain technology<sup>5</sup> |
| Tsang et al. | 2021 | J. Health. Eng. | Blockchain-IoT-driven nursing workforce planning for effective long-term care management in nursing homes<sup>6</sup> |
| Hughes | 2019 | PP Nursing P. | Blockchain and health care<sup>7</sup> |
| Niu | 2021 | J Health. Eng. | Influence of standardized nursing management of hospital based on smart electronic medical blockchain on nursing quality of digestive endoscopy room<sup>8</sup> |
| Anderson | 2018 | OJ Nursing Inf. | A look at blockchain and applicability to nursing<sup>9</sup> |
| Greenberger | 2019 | Nursing Mng. | Block what? the unrealized potential of blockchain in healthcare<sup>10</sup> |
| Li et al. | 2022 | Sci. Program. | Information Sharing and Privacy Protection of Electronic Nursing Record Management System<sup>11</sup> |

**Figure 1.**
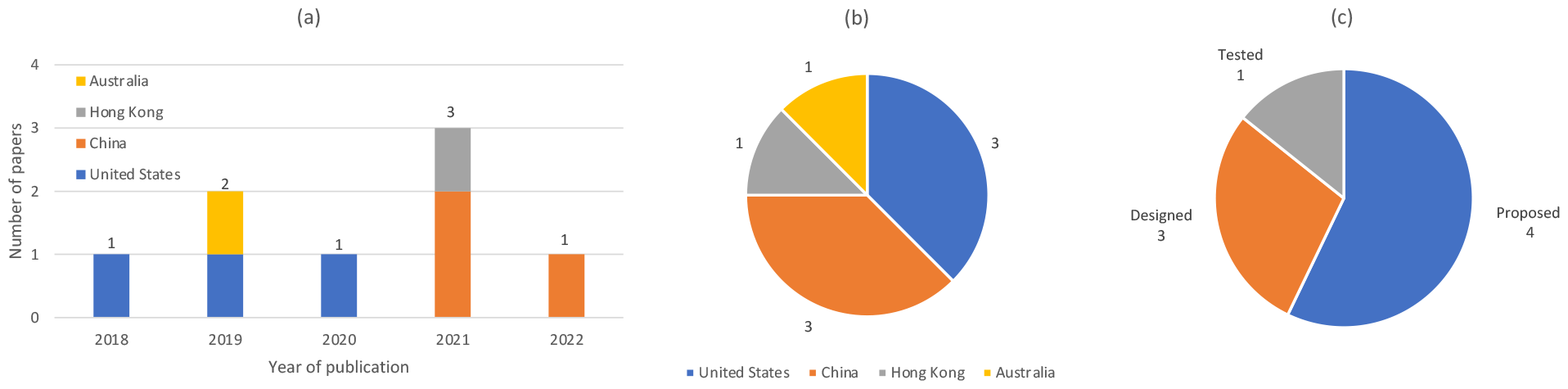
Key results by **(a)** publication years and countries, **(b)** countries only, and **(c)** blockchain adoption status.

## Discussion

We conducted a comparative study and extracted 13 data items from eight related articles, shedding light on the significant potential blockchain applications in nursing informatics across various domains. In recent years, there has been an increasing number of publications from Asia, indicating a growing research interest in exploring blockchain’s applications in nursing informatics. Integrating blockchain with innovative technologies is a promising approach to streamline processes, which can ultimately improve the quality of patient and healthcare outcomes. One limitation of our study is we are yet to identify more studies in languages other than English because the nursing blockchain applications are mostly in the early stages. Nevertheless, our research provides valuable insights into the potential of adopting blockchain into the nursing field for future nursing informatics researchers.

## Data Availability

All data produced in the present work are contained in the manuscript

## Acknowledgement

The authors were funded by the U.S. NIH (R00HG009680, R01EB031030, R01GM118609, R01HG011066, R01HL136835, RM1HG011558, T15LM011271, U24LM013755, and U54HG012510). The authors were also funded by the UC-Hispanic Serving Institutions Doctoral Diversity Initiative (UC-HSI DDI) and the UCSD Summer Training Academy for Research Success (STARS) program. The content is solely the responsibility of the authors and does not necessarily represent the official views of the NIH. The funders had no role in study design, data collection and analysis, decision to publish, or preparation of the manuscript.

## Notes

### Competing Interest Statement

The authors have declared no competing interest.

